# Characterization of a COPD-Associated *NPNT* Functional Splicing Genetic Variant in Human Lung Tissue via Long-Read Sequencing

**DOI:** 10.1101/2020.10.20.20203927

**Authors:** Aabida Saferali, Zhonghui Xu, Gloria M. Sheynkman, Craig P. Hersh, Michael H. Cho, Edwin K. Silverman, Alain Laederach, Christopher Vollmers, Peter J. Castaldi

## Abstract

Chronic obstructive pulmonary disease (COPD) is a leading cause of death worldwide. Genome-wide association studies (GWAS) have identified over 80 loci that are associated with COPD and emphysema, however for most of these loci the causal variant and gene are unknown. Here, we utilize lung splice quantitative trait loci (sQTL) data from the Genotype-Tissue Expression project (GTEx) and short read sequencing data from the Lung Tissue Research Consortium (LTRC) to characterize a locus in nephronectin (*NPNT*) associated with COPD case-control status and lung function. We found that the rs34712979 variant is associated with alternative splice junction use in *NPNT*, specifically for the junction connecting the 2nd and 4th exons (chr4:105898001-105927336) (p=4.02×10^−38^). This association colocalized with GWAS data for COPD and lung spirometry measures with a posterior probability of 94%, indicating that the same causal genetic variants in *NPNT* underlie the associations with COPD risk, spirometric measures of lung function, and splicing. Investigation of *NPNT* short read sequencing revealed that rs34712979 creates a cryptic splice acceptor site which results in the inclusion of a 3 nucleotide exon extension, coding for a serine residue near the N-terminus of the protein. Using Oxford Nanopore Technologies (ONT) long read sequencing we identified 13 *NPNT* isoforms, 6 of which are predicted to be protein coding. Two of these are full length isoforms which differ only in the 3 nucleotide exon extension whose occurrence differs by genotype. Overall, our data indicate that rs34712979 modulates COPD risk and lung function by creating a novel splice acceptor which results in the inclusion of a 3 nucelotide sequence coding for a serine in the nephronectin protein sequence. Our findings implicate *NPNT* splicing in contributing to COPD risk, and identify a novel serine insertion in the nephronectin protein that warrants further study.

## Introduction

The development of chronic obstructive pulmonary disease (COPD) is influenced by genetic susceptibility factors in addition to environmental exposures. Recent genome-wide association studies (GWAS) have identified over 80 distinct genetic loci that influence susceptibility to COPD and emphysema^1^, but for most of these loci the causal genetic variants and effector genes are unknown. By identifying the functional genetic variants in these GWAS loci and elucidating the biological mechanisms through which they influence disease susceptibility, causal mechanisms of COPD may be discovered.

The large majority of causal GWAS variants reside in the non-coding genome and disrupt gene regulatory elements. As a result, expression quantitative trait locus (eQTL) studies that associate genetic variants to gene expression values have been used to identify functional gene targets of GWAS-identified loci. In COPD, this approach has identified putative causal variants affecting the expression of *HHIP*^2^, *FAM13A*^3^, *TGFB2*^4^, and *ACVR1B*^5^. However, eQTL studies do not capture all of the potentially relevant functional mechanisms through which causal variants may alter gene expression. In particular, the alteration of gene splicing and isoform ratios is an important disease-causing gene regulatory mechanism that is not well captured by gene-level eQTL analyses^6,7^.

A genome-wide significant and replicated genetic association signal for respiratory phenotypes near *NPNT* may harbor a causal splicing variant, because the pattern of association at this locus is characterized by a single, clear lead SNP association that is located 5 nucleotides upstream from the 5’ splice site of the second exon in *NPNT*. We hypothesized that this region contains a functional variant that alters *NPNT* splicing. We utilized short and long-read RNA sequencing from human lung tissues to identify a causal splicing variant, rs34712979; to catalog the isoform variability of *NPNT* in the human lung; and to characterize the effects of rs34712979 on isoform usage rates.

## Methods

### Genetic association analysis, splicing QTL, and colocalization results

Summary genome-wide association study (GWAS) statistics were used from our previous study of COPD^1^ and two lung function phenotypes, FEV_1_ and FEV_1_/FVC (http://ldsc.broadinstitute.org/ldhub/)^8^. Leafcutter splicing QTL (sQTL) significant results^9^ were obtained for all tissues from the GTEx Portal (https://www.gtexportal.org/home/datasets), and complete lung sQTL results were obtained from the Anvil GTEx Terra workspace. Multiple colocalization for GWAS and sQTL results was performed using the moloc R package (https://github.com/clagiamba/moloc). Moloc analyses were performed using default parameters for prior variance of the approximate Bayes factor (ABF, prior_var = c(0.01, 0.1, 0.5)) and default parameters for the prior likelihood that a given SNP is causal for one trait, pairs of traits, or all traits (priors = c(1e-04, 1e-06, 1e-07). Linkage disequilibrium (LD), evolutionary conservation, and overlap with regulatory elements were identified for individual single nucleotide polymorphisms (SNPs) with Haploreg (https://pubs.broadinstitute.org/mammals/haploreg/haploreg.php). Posterior causal probabilities based on strength of genetic association and local LD patterns were determined using the PICS algorithm (https://pubs.broadinstitute.org/pubs/finemapping/pics.php).^10^

### Splicing analysis in short read RNA-seq data from GTEx lung tissue samples

With dbGaP approval, RNA-seq BAM files and whole genome sequencing VCF files for lung tissue samples in GTEx V8 release were accessed on Google Cloud via the AnVIL GTEx Terra workspace (https://app.terra.bio/#workspaces/anvil-datastorage/AnVIL_GTEx_V8_hg38). To obtain splicing junctional counts for each genotype and visualize with Sashimi plots, a Docker image (https://hub.docker.com/repository/docker/pacifly/splice-plot-app) was built on the Python package SplicePlot^11^ (https://github.com/wueric/SplicePlot) and its dependencies, and a WDL workflow (https://portal.firecloud.org/?return=terra#methods/zxu_spliceplot/spliceplot/18) was created and executed via Cromwell engine to run SplicePlot functionalities on Google Cloud. The splicing junctional count method in SplicePlot was adapted to accommodate novel junctions missed from the alignment.

### Lung Tissue Research Consortium Samples, short-read RNA sequencing, and whole genome sequencing

The Lung Tissue Research Consortium (LTRC) is an NHLBI-sponsored collection of lung and blood tissues collected from patients undergoing thoracic surgery who completed a standard questionnaire, pulmonary function testing, and chest computed tomography (CT) imaging. Through the NHLBI Trans-Omics and Precision Medicine (TOPMed) program, LTRC whole-genome sequencing (WGS) data were generated at Broad Genomics and lung RNA-seq were generated at the University of Washington, and 1,335 LTRC samples with RNA-seq and WGS passing quality control filters were analyzed in this study. Briefly, RNA-seq data were generated using paired-end Illumina sequencing from poly-A selected libraries to an average depth of 67 million mapped reads. Methods for TOPMed WGS are available at https://www.nhlbiwgs.org/topmed-whole-genome-sequencing-methods-freeze-8.

### Long read RNA-seq analysis in human lung samples from the LTRC

We conducted targeted Oxford Nanopore Technologies (ONT) long read sequencing on RNA from 10 human lung samples from the LTRC which were selected to include five samples from each homozygous class (i.e., GG genotype, AA genotype) of rs34712979. For each of these ten samples, 100-200ng of total RNA was used to generate full-length cDNA using a modified Smart-seq2 protocol^12^. The enrichment and library generation procedures are described in detail in the Supplemental Methods.

Enriched re-amplified cDNA from two independent enrichment reactions was sequenced on a MinION 9.4.1 flow cell or a MinION 10.3 flow cell using the R2C2 method^13-16^. For each run, 1ug of DNA was prepared using the LSK-109 kit according to the manufacturer’s instructions with only minor modifications. End-repair and A-tailing steps were both extended from 5 minutes to 30 minutes. The final ligation step was also extended to 30 minutes. Each run took 48 hours and the resulting data in Fast5 format was basecalled using the high accuracy model of the gpu accelerated Guppy algorithm (9.4.1 flow cell: version 3.4.5+fb1fbfb with config file dna_r9.4.1_450bps_hac.cfg config file; 10.3 flow cell: version 3.6.1+249406c with config file dna_r10_450bps_hac.cfg). To generate R2C2 consensus reads for each sample, we processed and demultiplexed the resulting raw reads using our C3POa pipeline (https://github.com/rvolden/C3POa). R2C2 reads were analyzed to identify and quantify isoforms using version 3.5 of Mandalorion (-O 0,40,0,40 -r 0.01 -i 1 -w 1 -n 2 -R 5) (https://github.com/rvolden/Mandalorion-Episode-III). Isoforms were categorized using the sqanti_qc.py script of the SQANTI^17^ program with slight modifications to make it compatible with Python3.

For *NPNT* isoform quantification (i.e., usage analysis), the proportion of isoform usage for each sample was calculated by dividing the number of reads for each isoform by the total number of isoform reads aligning to the *NPNT* locus. Differences in isoform usage between genotype classes were identified using the Mann-Whitney test. For certain analyses, isoform reads were collapsed based on whether they contained a 3 nucleotide exon extension. To quantify the number of reads containing this TAG sequence at the 5’ end of the second exon, we extraEdwin K. Silverman, MD, PhDc ted the following 30-mers from the fasta files for each sample using the ‘grep’ command:

> AGTTCGACGGGAG<u>TAG</u>GTGGCCCAGGCAAA
>
> CGAGTTCGACGGGAGGTGGCCCAGGCAAAT

### NPNT protein sequence analysis

Protein sequence analysis was performed using Uniprot^18^ to identify NPNT protein domains. The Chou and Fasman Secondary Structure Prediction server^19^ was used to characterize the impact of sequence changes to NPNT structure.

## Results

### Genetic association signals for respiratory phenotypes in NPNT

Genome-wide significant association signals near *NPNT* have been identified for COPD and various measures of lung function, and previous colocalization and fine mapping analyses have implicated rs34712979 as the most likely causal variant for the COPD association near *NPNT*^1^. We obtained the summary GWAS statistics for the most recent GWAS meta-analyses of COPD, FEV_1_, and FEV_1_/FVC and compared the genetic association signal for each of these phenotypes. In each case, the variant with minimal p-value was rs34712979, a common variant with a minor allele frequency of 23% in the 1,000 Genomes European (EUR) population. Using the PICS algorithm, we determined that the estimated posterior probability that rs34712979 was the causal variant for each of these associations was between 94-100%. This variant is in strong linkage disequilibrium with only one other common variant, rs6828309 (r^2^ =0.81 in EUR). Supplemental Table 1 shows that rs34712979 is highly conserved and overlaps promoter and enhancer elements in multiple cell types, which is not the case for rs6828309 (Supplemental Table 1). The minor A allele of rs34712979 is associated with lower measures of lung function and increased risk of COPD (odds ratio=1.18).

### An NPNT splicing association signal in human lung tissue colocalizes with GWAS associations

To determine whether the genetic associations near *NPNT* to pulmonary phenotypes may be explained by genetic effects on splicing, we examined leafcutter quantitative trait locus (QTL) results from 49 tissues obtained from a total of 838 subjects in GTEx version 8. We observed that rs34712979 was associated with splicing ratios for three exon-exon junctions in a total of 18 tissues that connect the first and second, second and fourth, and third and fourth exons of *NPNT* (GENCODE v32) (Supplemental Table 2). Focusing on lung tissue, the most relevant tissue for COPD, we observed that rs34712979 was associated with multiple splicing ratios at a nominal p-value threshold of p<0.001 (Supplemental Table 3), with the association for the junction connecting the 2nd and 4th exons (chr4:105898001-105927336) exceeding genome-wide significance. To better understand the effect of rs34712979 on splicing in human lung, we identified 27 subjects in GTEx homozygous for the A allele of rs34712979 and compared the distribution of junctional reads from lung RNA in these subjects against 27 randomly selected samples from the other two genotype classes (Figure 1), and found that the A allele results in higher rates of inclusion of the 3rd exon. Upon further investigation of junctional reads flanking exon 2 we discovered the presence of a novel splice site acceptor at the 5’ end of exon 2 associated with the A allele. This novel cryptic splice site was not detected in the sQTL analysis as 99.6% of reads using the cryptic splice site were soft-clipped (ie. portions of the read were masked due to a mismatch with the reference genome) and therefore were not included in the Leafcutter clustering step.

**Figure 1.**
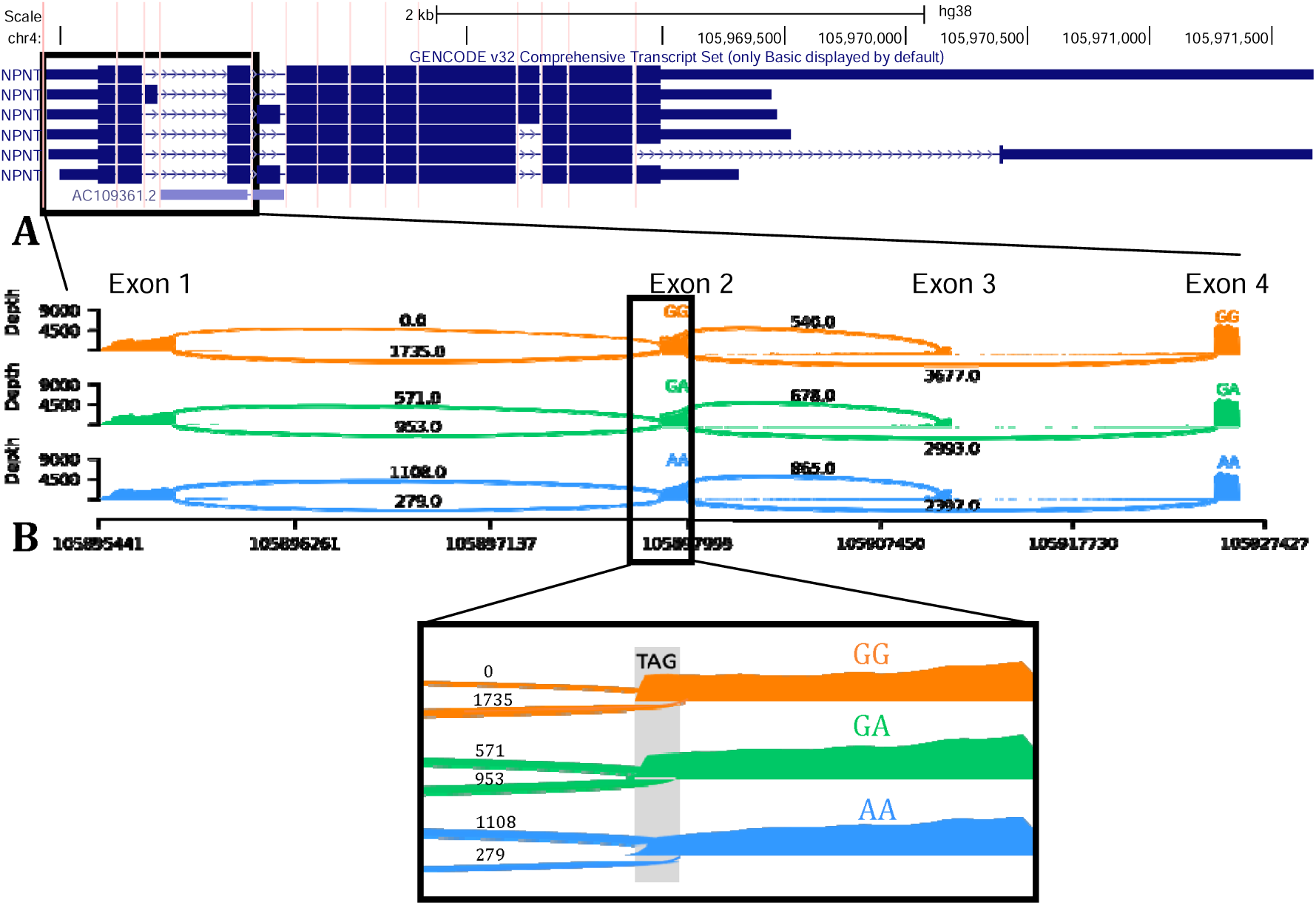
GTEx Leafcutter lung sQTL results for effect of rs34712979 on splice junctions involving the 2^nd^-4^th^ exons of *NPNT* in leafcutter splicing analysis from short-read RNAseq in 515 GTEx samples (A) and in junctional reads from 27 samples from each genotype class of rs34712979 (B). The 3 nucleotide alternatively spliced exonic extension sequence is shown in the inset window of panel B.

To confirm that the lung sQTL signals in this region overlap with the GWAS association signals, we performed multiple colocalization using the moloc method which resulted in an estimated 94% probability of a shared causal variant for the three genetic association signals and the lung splicing signal for the 2nd and 4th exons of *NPNT* (chr4:105898001-105927336). rs34712979 had the best individual SNP posterior probability across all evaluated scenarios, with a posterior probability of 94% for the scenario of shared colocalization across all four datasets (Supplemental Table 4). The local association plots for each of the association signals near *NPNT* is shown in Figure 2.

**Figure 2.**
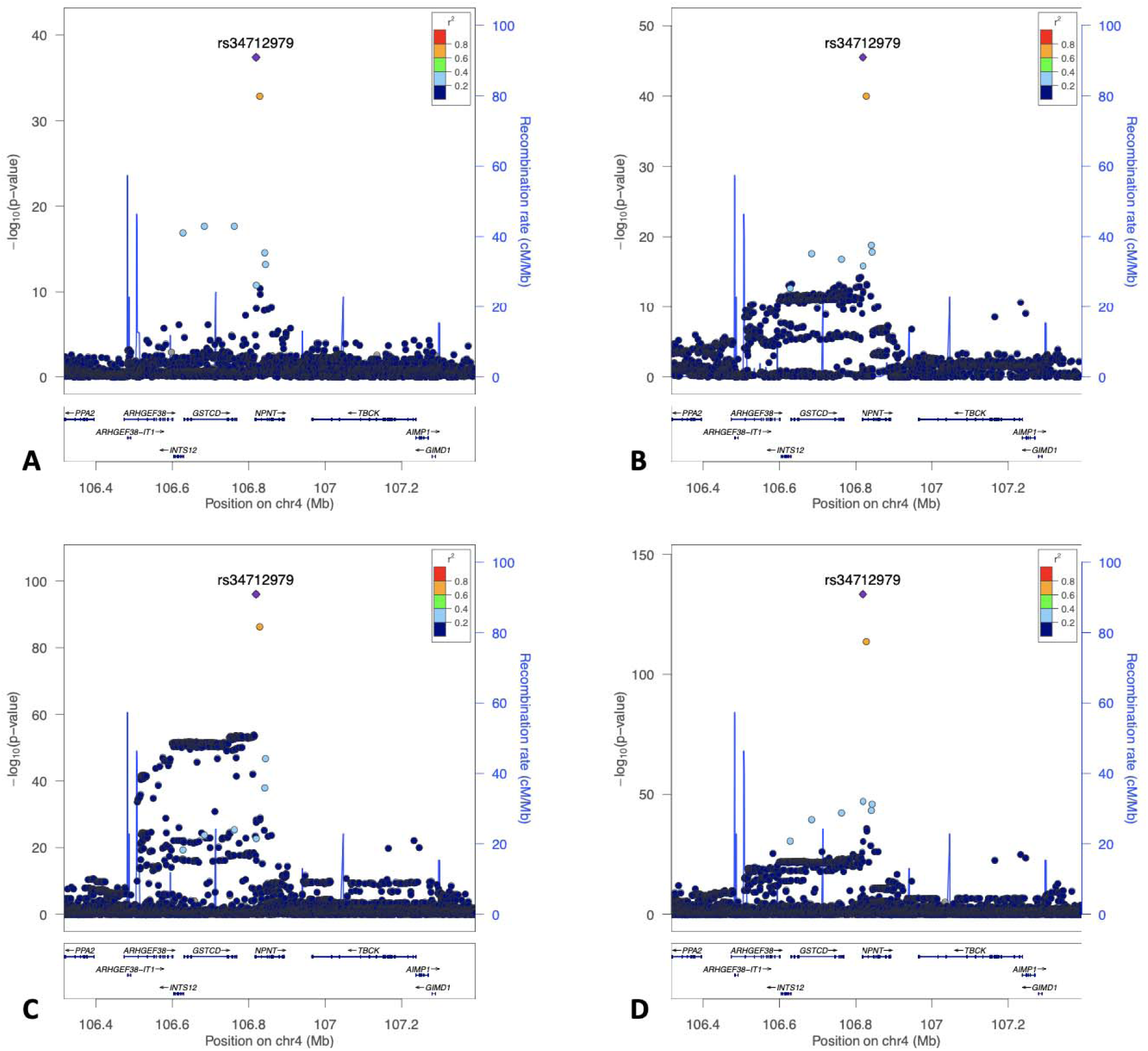
Local association plots for genetic association near *NPNT* to COPD (A, Sakornsakolpat 2019), FEV1 (B), FEV1/FVC (C, Shrine 2019), and GTEx Leafcutter lung sQTL analyses (D).

### rs34712979 creates an alternative splice acceptor site

rs34712979 is located 5bp upstream of the second exon of *NPNT*, so we examined the sequence content in this region for motifs that may alter splicing, and we observed that the minor A allele of rs34712979 creates a cryptic AG splice acceptor that would result in a 3 nucleotide 5’ exon extension in exon 2. In other words, the A allele creates a NAGNAG splice site, which contains adjacent AG acceptor sites that can be variably used^20,21^. Open reading frame analysis indicates that this results in an additional in-frame AGT codon, coding for serine, spanning the boundaries of the first and second exon (Figure 3).

**Figure 3.**
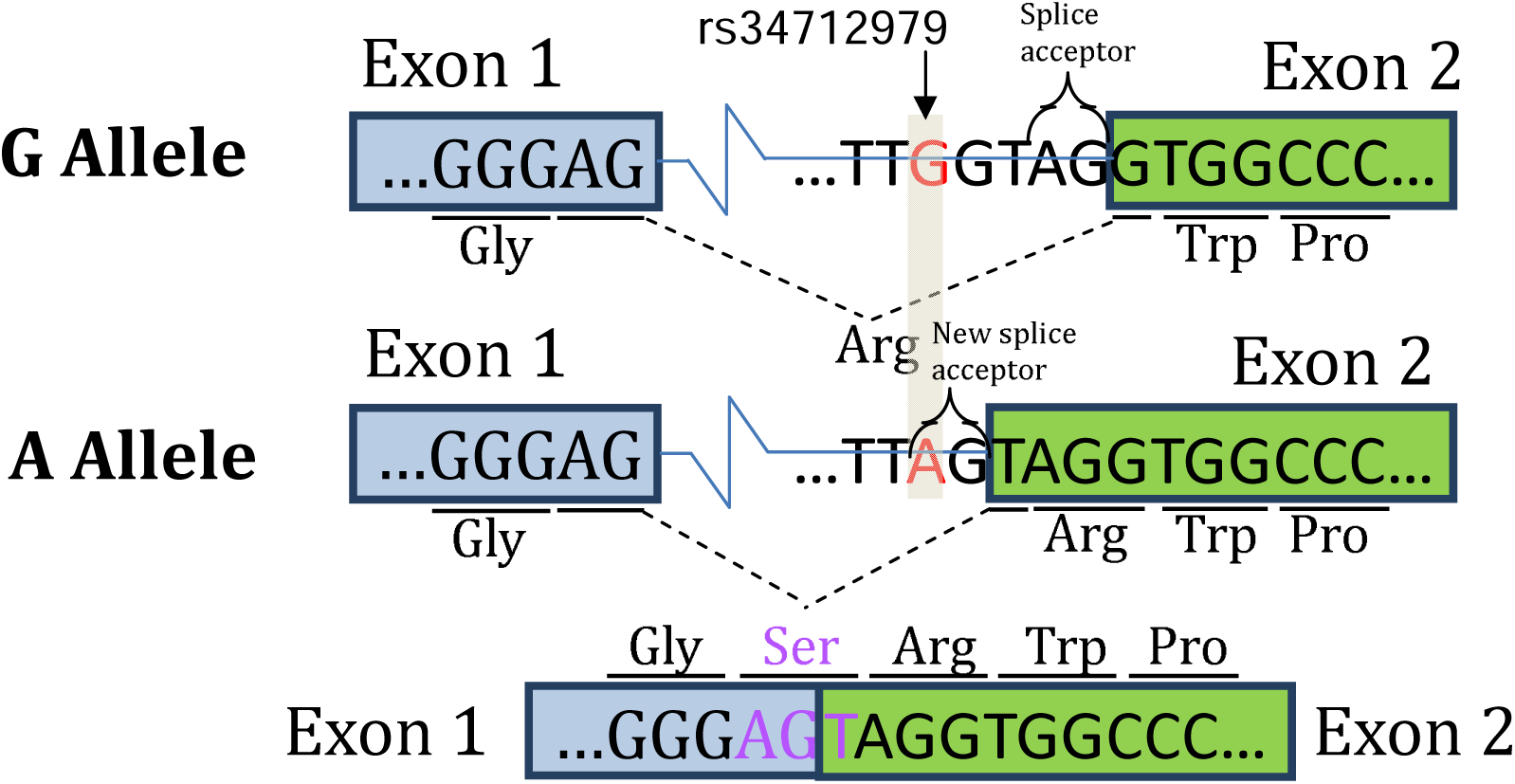
The A allele of rs34712979 creates an alternative splice acceptor site resulting in inclusion of a serine at the 5’ splice site of exon 2.

To confirm this effect in human lung samples, we queried short-read RNA-seq data from human lung samples in GTEx and confirmed that samples with the A allele demonstrate preferential use of the cryptic versus the annotated splice acceptor site (Figure 4). We further confirmed that this phenomenon occurs in a larger set of 1,335 lung samples from the LTRC (Figure 4).

**Figure 4.**
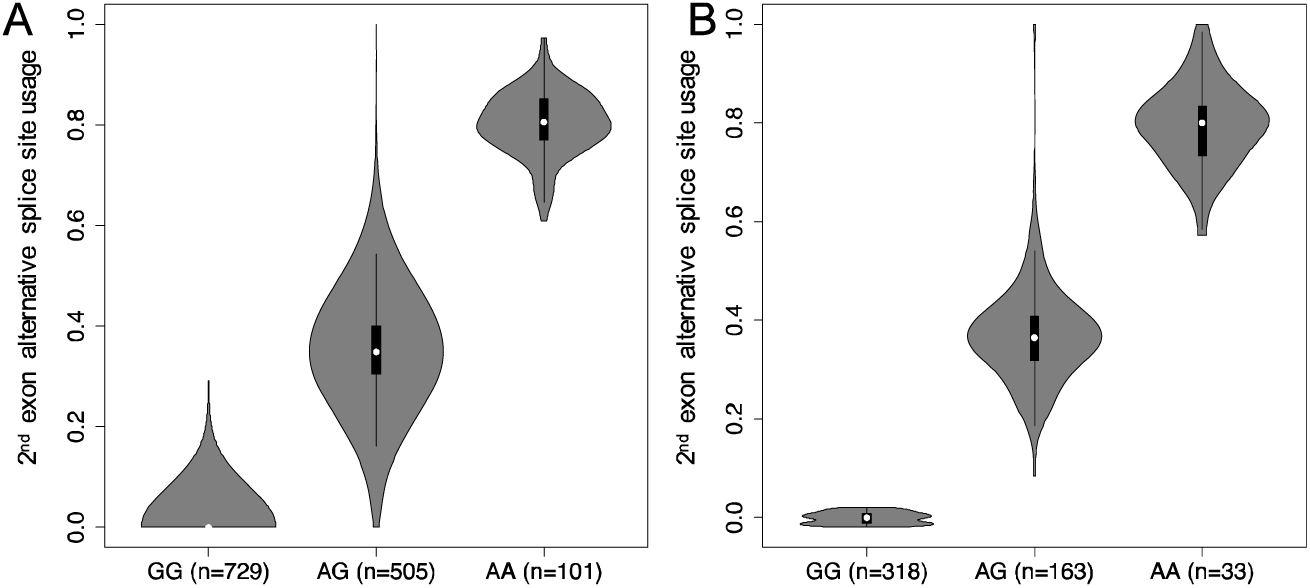
Violin plot of exon 2 alternative splice site usage in short-read RNA-seq from A) LTRC and B) GTEX Lung Tissue Samples.

### Characterization of rs34712979 Allelic Effects on NPNT Isoform Usage

To determine the effect of rs34712979 on full-length NPNT isoforms, we performed targeted enrichment for *NPNT* transcripts followed by ONT long-read sequencing in lung tissue samples from the LTRC selected to include 5 subjects from each homozygote class of rs34712979. The experiment yielded 24,747 reads mapping to *NPNT*. Thirteen high confidence isoforms were detected, 12 of which were novel (Supplemental Table 1). Six of these isoforms, including one antisense isoform, have open reading frames indicating that they have protein-coding potential. Two protein coding isoforms containing exon 2 utilize the cryptic splice acceptor, while two isoforms utilize the annotated splice acceptor (Figure 5, panel A).

**Figure 5.**
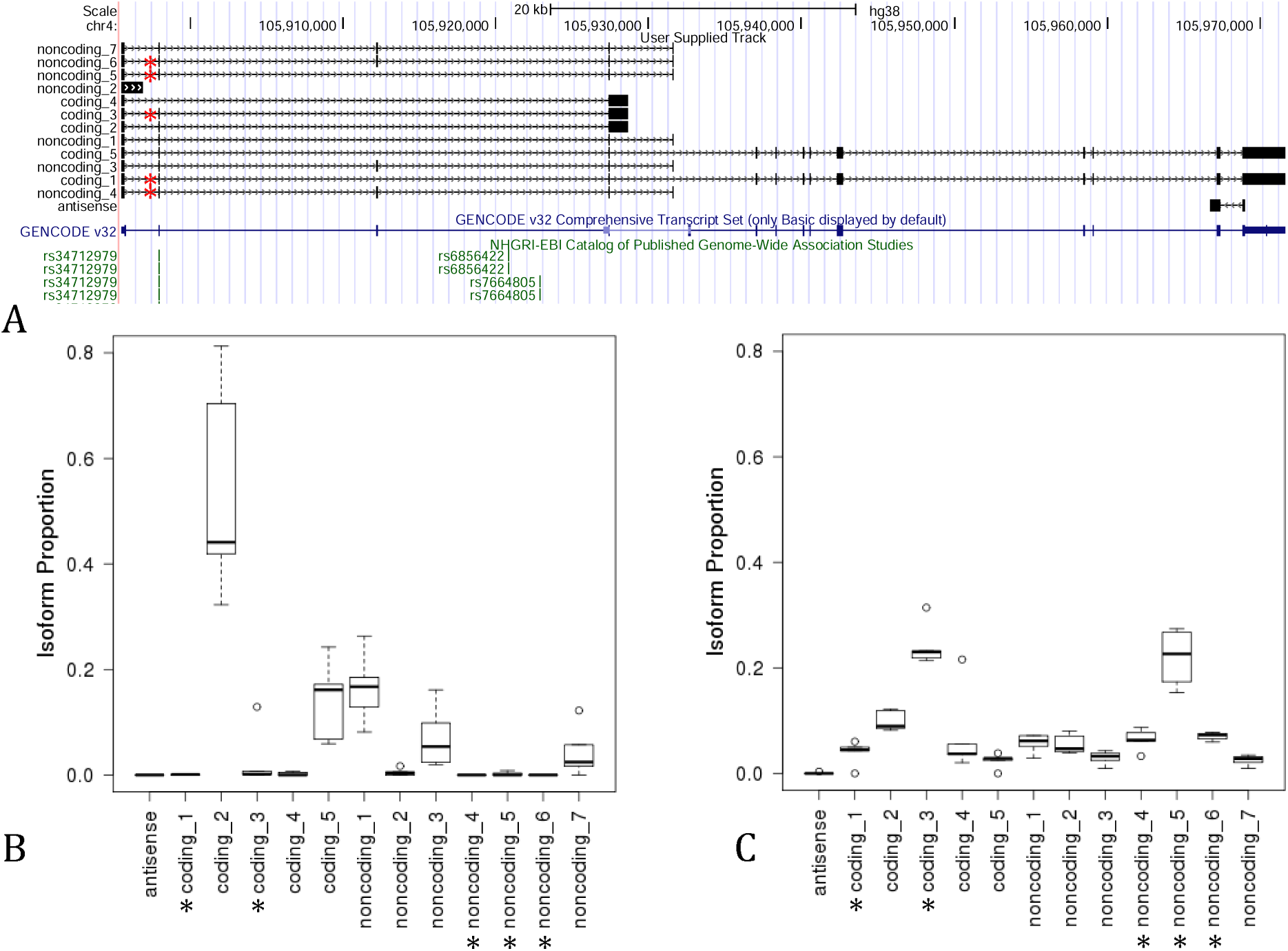
Isoforms detected by long read sequencing in 10 human lung tissues predicted to be coding or noncoding (A) * Indicates isoforms utilizing the novel splice acceptor site resulting in a three nucleotide 5’ extension of exon 2. The isoform usage profile differs markedly between rs34712979 GG (B) and AA homozygotes (C).

Isoform usage differed between rs34712979 genotype classes. On a per-isoform basis, 9 of the 13 isoforms show differential usage between the two homozygous genotype classes (Mann-Whitney p<0.05). Focusing on the putative protein-coding isoforms, we found that the full length annotated isoform ‘coding_5’ (corresponding to ENST00000379987.6) is more highly expressed in the GG genotype class, while isoform ‘coding_1’, also full length, is highly expressed in 4 out of 5 subjects with the AA genotype (Figure 6). The three short isoforms are also differentially expressed by genotype, with isoforms ‘coding 3’ and ‘coding 4’ more highly expressed in AA. Looking specifically at usage of the alternative or canonical splice acceptor site at the 5’ end of exon 2, we observed 10 isoforms that include the second exon, with five isoforms each using the alternative or canonical splice acceptor. Collapsing these isoforms by splice site usage demonstrates markedly increased usage of the alternative splice site in the AA genotype, with essentially no usage of the alternative site in 4 of the 5 GG genotype class samples (Figure 6, panel E). We confirmed the presence of the 3-nucleotide 5’ exon extension sequence in full length reads by counting the reads containing a 30-bp sequence centered on the TAG triplet located after the novel splice junction and compared this to the number of reads containing a corresponding 30-bp sequence without the TAG. Table 1 shows the clear association between the AA genotype and the 3-nucleotide exon extension event.

**Table 1.** Number of ONT reads containing 30bp sequences including and excluding the TAG sequence at the 5’ end of the 2nd exon of NPNT.

| rs34712979 | ...GATTAGGTG... | ...GATGTG... |
| --- | --- | --- |
| AA | 825 | 206 |
| AA | 735 | 163 |
| AA | 291 | 92 |
| AA | 1638 | 403 |
| AA | 2041 | 500 |
| GG | 10 | 2216 |
| GG | 2 | 1519 |
| GG | 8 | 51 |
| GG | 6 | 502 |
| GG | 1 | 1553 |

**Figure 6.**
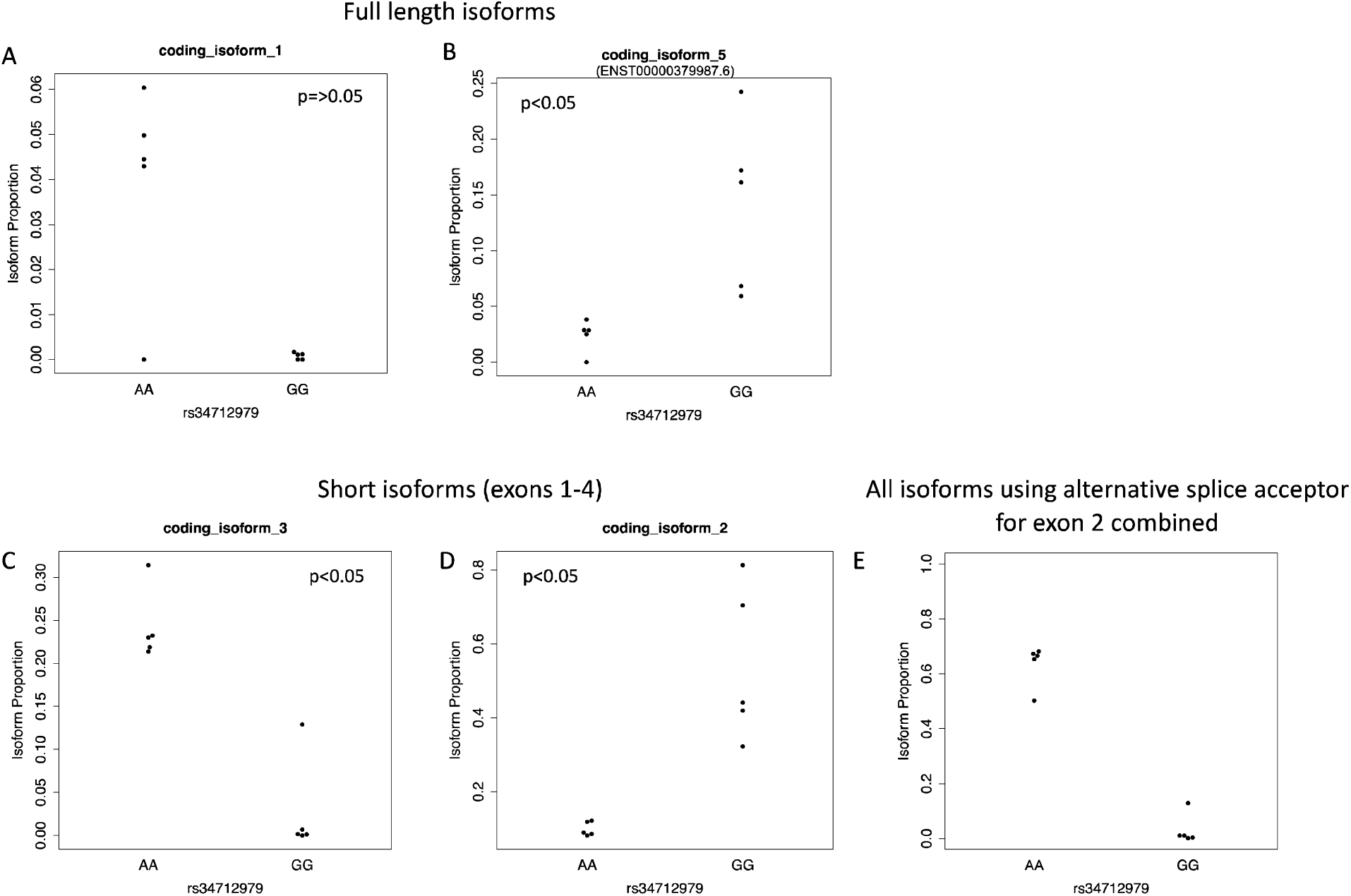
Long read sequencing confirms the presence of two full length isoforms, one incorporating a 3 bp intronic sequence (A) and the other which is fully annotated (B). Out of the two short isoforms (coding isoforms 2 and 3) which contain exon 2, one includes the upstream cryptic splice site and is more highly expressed in samples homozygous for the rs34712979 AA genotype (C), while the other is predominantly expressed in the GG genotype (D). All isoforms using the cryptic splice site combined have increased expression in the AA genotype with minimal expression in GG.

### Protein sequence analysis

The serine residue 5’ exon extension is inserted 24 residues from the N-terminus of NPNT protein, and four amino-acids after the end of the predicted signal peptide. Protein secondary structure analysis of NPNT isoform sequences with and without the 3 nucleotide exon extension revealed that the serine residue results in the perturbation of an alpha-helical segment with a turn motif (Supplemental Figure 1).

## Discussion

Our results provide strong support for the hypothesis that rs34712979 is a functional GWAS variant that acts by altering isoform usage of *NPNT*. Our analyses led to three main findings: 1) the genetic association patterns of association to COPD, FEV1 and FEV1/FVC near *NPNT* are highly likely to share rs34712979 as a causal genetic variant, 2) the A allele of rs34712979 creates an alternative splice acceptor site that results in a 3-nucleotide exon extension at the 5’ end of exon 2 that is predicted to result in the addition of a serine to the NPNT protein, and 3) the A allele alters *NPNT* isoform usage. The genetic association patterns reflect the cumulative data of hundreds of thousands of subjects, and the splicing effects were demonstrated in lung RNA from over 1,000 subjects in two different human cohorts.

NPNT, or nephronectin, is an extracellular protein involved in tissue development, remodeling, and repair. It was first identified in the context of the search for a novel ligand for integrin α8β1^22,23^and was shown to be necessary for normal kidney development in murine models^24^. NPNT has also been linked to osteoblast differentiation and bone remodeling^25^, invasiveness of breast cancer^26^, and pulmonary silicosis^27^. Full-length NPNT protein includes five epidermal growth factor (EGF)-like functional domains followed by a meprin, A-5 protein, receptor protein-tyrosine phosphatase mu (MAM), and an Arg-Gly-Asp (RGD) integrin-binding domain. The most well-described function of NPNT is as an extra-cellular matrix protein that binds integrin α8β1 through its RGD domain. Recently, NPNT has been identified in extracellular vesicles secreted by a murine breast cancer cell line, and full-length NPNT has been reported to undergo post-translational modifications including cleavage to a shorter 20-kd isoform^28^. Full length NPNT has also been shown to be highly expressed in human pneumocytes in the Human Protein Atlas project^29^. Previous studies have identified NPNT as a potential effector gene via association between lung function and mRNA levels and NPNT staining in pulmonary endothelial and alveolar epithelial cells^30^. In our study, we identify two full length NPNT isoforms, one of which is fully annotated while the other utilizes a cryptic splice acceptor in exon 2, in addition to three shortened isoforms that are likely protein coding. Within the pair of long isoforms and within the pair of short isoforms, they differ only in the inclusion of a three nucleotide 5’ exon extension coding for a serine residue near the N-terminus of the protein. Additional studies are needed to confirm the extent to which these differences at the RNA level translate to differences in pulmonary protein isoform content, structure and function.

The *NPNT* genetic association lies in a region first described in association with lung function^31,32^. Subsequent studies confirmed an association with COPD and also identified two independent signals at this locus^33^. Several genes in this region have previously been hypothesized to be the effector genes using eQTL studies, including the nearby genes *GSTCD* and *INTS12*. These two genes harbor eQTL in blood and lung, and prior studies have found correlations between *GSTCD* and *INTS12* mRNA with lung function and variable expression during lung development^34^. Additional studies identified *NPNT* as another potential effector gene at this locus^1,30^, indicating that the genetic association at this locus likely involves several different genes and mechanisms.

From a genetic standpoint, the most clear and compelling genetic associations of *NPNT* to common disease colocalize to an area including the second exon with clear evidence that rs34712979 is a causal variant contributing to the association signals to lung function^8,35^ and COPD^1^. Our RNA-seq analyses provide strong evidence that the A allele of rs34712979 (which is associated with decreased pulmonary function and increased COPD risk) creates an alternative splice acceptor site at the 5’ end of the second exon, and this site is preferentially used relative to the annotated splice site on haplotypes containing the A allele. Usage of this alternative splice site results in the inclusion of a serine residue near the N-terminus of the protein that is predicted to perturb an alpha-helical segment with a turn motif. In addition to this alternative acceptor site usage, the A allele substantially alters *NPNT* isoform usage patterns. While some of the isoform changes can be directly explained by the second exon alternative splice site usage event, our analysis also identified allelic effects on downstream splicing events as well. The mechanisms that may link usage of alternative splice site to other splicing events remain to be elucidated.

As is characteristic of human GWAS associations, the penetrance of the rs34712979 A allele is low, indicating that either the overall effect of this allele on biological function is subtle or that the effect is large but occurs only in restricted sub-populations. Additional research on *NPNT* isoform expression at the RNA and protein levels in large, well-phenotyped human cohorts may identify specific sub-populations most impacted by the functional consequences of this allele.

The strengths of this study are that the GWAS and RNA-seq findings are based on the analysis of a very large number of human samples, and the integrated analysis of short and long-read RNA-seq data provides a high level of resolution to observe changes at the isoform level.

To our knowledge this is one of the first applications of long-read sequencing to human tissue samples to demonstrate splicing-related effects of a GWAS-identified genetic variant. Important limitations to consider are that long-read sequencing technologies can be affected by biases related to transcript length, therefore direct comparisons of abundance between the short and long isoforms of *NPNT* should be interpreted with caution. The most prominent finding in our study, namely allele-specific alternative splice site usage, is clearly identified in isoforms of equal length suggesting that these allele-specific observations are unlikely to be affected by this bias. While there is strong statistical support for our findings, these are nonetheless correlative findings from large human cohorts and future work is required to define the underlying molecular mechanisms and provide further experimental evidence to demonstrate how these mechanisms may alter COPD risk.

In summary, these analyses demonstrate that the pulmonary disease GWAS association near NPNT is very likely to be mediated by a common genetic variant that alters splicing, resulting in the insertion of a novel serine residue in the NPNT protein immediately downstream of the signal peptide domain. Given the known function of NPNT in extracellular matrix biology and the high expression of NPNT in lung tissue and pneumocytes^30^, further investigation of the functional consequences of this splicing variant, including its effects on NPNT protein structure and function, are likely to elucidate causal mechanisms of COPD pathogenesis.

## Data Availability

All data will be publicly available through dbGaP (accession number phs001662.v1.p1)

## Funding/Acknowledgements

This work was funded by R01 HL124233, R01 HL147326, R01 HL111527, U01 HL089897, U01 HL089856, R01HL125583, R01HL130512, T32HL007427. Research reported in this publication was supported by the NHLBI and FDA Center for Tobacco Products (CTP). The content is solely the responsibility of the authors and does not necessarily represent the official views of the NIH or the Food and Drug Administration.

The Genotype-Tissue Expression (GTEx) Project was supported by the <u>Common Fund</u> of the Office of the Director of the National Institutes of Health, and by NCI, NHGRI, NHLBI, NIDA, NIMH, and NINDS. The data used for the analyses described in this manuscript were obtained from: the GTEx Portal between January and August of 2020 and via the GTEx Terra Workspace during the same time interval.

Molecular data for the Trans-Omics in Precision Medicine (TOPMed) program was supported by the National Heart, Lung and Blood Institute (NHLBI). Whole Genome Sequencing and RNASeq for “NHLBI TOPMed: The Lung Tissue Research Consortium (phs001662)” was performed at Northwest Genome Center (NWGC, HHSN268201600032I, RNASeq) and Broad Genomics (HHSN268201600034I, WGS) Core support including centralized genomic read mapping and genotype calling, along with variant quality metrics and filtering were provided by the TOPMed Informatics Research Center (3R01HL-117626-02S1; contract HHSN268201800002I). Core support including phenotype harmonization, data management, sample-identity QC, and general program coordination were provided by the TOPMed Data Coordinating Center (R01HL-120393; U01HL-120393; contract HHSN268201800001I). We gratefully acknowledge the studies and participants who provided biological samples and data for TOPMed.

## COPDGene^®^ Investigators – Core Units

*Administrative Center*: James D. Crapo, MD (PI); Edwin K. Silverman, MD, PhD (PI); Barry J. Make, MD; Elizabeth A. Regan, MD, PhD

*Genetic Analysis Center*: Terri Beaty, PhD; Ferdouse Begum, PhD; Peter J. Castaldi, MD, MSc; Michael Cho, MD; Dawn L. DeMeo, MD, MPH; Adel R. Boueiz, MD; Marilyn G. Foreman, MD, MS; Eitan Halper-Stromberg; Lystra P. Hayden, MD, MMSc; Craig P. Hersh, MD, MPH; Jacqueline Hetmanski, MS, MPH; Brian D. Hobbs, MD; John E. Hokanson, MPH, PhD; Nan Laird, PhD; Christoph Lange, PhD; Sharon M. Lutz, PhD; Merry-Lynn McDonald, PhD; Margaret M. Parker, PhD; Dmitry Prokopenko, Ph.D; Dandi Qiao, PhD; Elizabeth A. Regan, MD, PhD; Phuwanat Sakornsakolpat, MD; Edwin K. Silverman, MD, PhD; Emily S. Wan, MD; Sungho Won, PhD

*Imaging Center*: Juan Pablo Centeno; Jean-Paul Charbonnier, PhD; Harvey O. Coxson, PhD; Craig J. Galban, PhD; MeiLan K. Han, MD, MS; Eric A. Hoffman, Stephen Humphries, PhD; Francine L. Jacobson, MD, MPH; Philip F. Judy, PhD; Ella A. Kazerooni, MD; Alex Kluiber; David A. Lynch, MB; Pietro Nardelli, PhD; John D. Newell, Jr., MD; Aleena Notary; Andrea Oh, MD; Elizabeth A. Regan, MD, PhD; James C. Ross, PhD; Raul San Jose Estepar, PhD; Joyce Schroeder, MD; Jered Sieren; Berend C. Stoel, PhD; Juerg Tschirren, PhD; Edwin Van Beek, MD, PhD; Bram van Ginneken, PhD; Eva van Rikxoort, PhD; Gonzalo Vegas Sanchez-Ferrero, PhD; Lucas Veitel; George R. Washko, MD; Carla G. Wilson, MS;

*PFT QA Center, Salt Lake City, UT*: Robert Jensen, PhD

*Data Coordinating Center and Biostatistics, National Jewish Health, Denver, CO*: Douglas Everett, PhD; Jim Crooks, PhD; Katherine Pratte, PhD; Matt Strand, PhD; Carla G. Wilson, MS

*Epidemiology Core, University of Colorado Anschutz Medical Campus, Aurora, CO*: John E. Hokanson, MPH, PhD; Gregory Kinney, MPH, PhD; Sharon M. Lutz, PhD; Kendra A. Young, PhD

*Mortality Adjudication Core:* Surya P. Bhatt, MD; Jessica Bon, MD; Alejandro A. Diaz, MD, MPH; MeiLan K. Han, MD, MS; Barry Make, MD; Susan Murray, ScD; Elizabeth Regan, MD; Xavier Soler, MD; Carla G. Wilson, MS

*Biomarker Core*: Russell P. Bowler, MD, PhD; Katerina Kechris, PhD; Farnoush Banaei-Kashani, Ph.D **COPDGene**^**®**^ **Investigators – Clinical Centers**

*Ann Arbor VA:* Jeffrey L. Curtis, MD; Perry G. Pernicano, MD

*Baylor College of Medicine, Houston, TX*: Nicola Hanania, MD, MS; Mustafa Atik, MD; Aladin Boriek, PhD; Kalpatha Guntupalli, MD; Elizabeth Guy, MD; Amit Parulekar, MD;

*Brigham and Women’s Hospital, Boston, MA*: Dawn L. DeMeo, MD, MPH; Alejandro A. Diaz, MD, MPH; Lystra P. Hayden, MD; Brian D. Hobbs, MD; Craig Hersh, MD, MPH; Francine L. Jacobson, MD, MPH; George Washko, MD

*Columbia University, New York, NY*: R. Graham Barr, MD, DrPH; John Austin, MD; Belinda D’Souza, MD; Byron Thomashow, MD

*Duke University Medical Center, Durham, NC*: Neil MacIntyre, Jr., MD; H. Page McAdams, MD; Lacey Washington, MD

*Grady Memorial Hospital, Atlanta, GA: Eric Flenaugh, MD; Silanth Terpenning, MD HealthPartners Research Institute, Minneapolis, MN*: Charlene McEvoy, MD, MPH; Joseph Tashjian, MD

*Johns Hopkins University, Baltimore, MD*: Robert Wise, MD; Robert Brown, MD; Nadia N. Hansel, MD, MPH; Karen Horton, MD; Allison Lambert, MD, MHS; Nirupama Putcha, MD, MHS

*Lundquist Institute for Biomedical Innovationat Harbor UCLA Medical Center, Torrance, CA*: Richard Casaburi, PhD, MD; Alessandra Adami, PhD; Matthew Budoff, MD; Hans Fischer, MD; Janos Porszasz, MD, PhD; Harry Rossiter, PhD; William Stringer, MD

*Michael E. DeBakey VAMC, Houston, TX*: Amir Sharafkhaneh, MD, PhD; Charlie Lan, DO

*Minneapolis VA:* Christine Wendt, MD; Brian Bell, MD; Ken M. Kunisaki, MD, MS

*National Jewish Health, Denver, CO*: Russell Bowler, MD, PhD; David A. Lynch, MB

*Reliant Medical Group, Worcester, MA*: Richard Rosiello, MD; David Pace, MD

*Temple University, Philadelphia, PA:* Gerard Criner, MD; David Ciccolella, MD; Francis Cordova, MD; Chandra Dass, MD; Gilbert D’Alonzo, DO; Parag Desai, MD; Michael Jacobs, PharmD; Steven Kelsen, MD, PhD; Victor Kim, MD; A. James Mamary, MD; Nathaniel Marchetti, DO; Aditi Satti, MD; Kartik Shenoy, MD; Robert M. Steiner, MD; Alex Swift, MD; Irene Swift, MD; Maria Elena Vega-Sanchez, MD

*University of Alabama, Birmingham, AL:* Mark Dransfield, MD; William Bailey, MD; Surya P. Bhatt, MD; Anand Iyer, MD; Hrudaya Nath, MD; J. Michael Wells, MD

*University of California, San Diego, CA*: Douglas Conrad, MD; Xavier Soler, MD, PhD; Andrew Yen, MD

*University of Iowa, Iowa City, IA*: Alejandro P. Comellas, MD; Karin F. Hoth, PhD; John Newell, Jr., MD; Brad Thompson, MD

*University of Michigan, Ann Arbor, MI:* MeiLan K. Han, MD MS; Ella Kazerooni, MD MS; Wassim Labaki, MD MS; Craig Galban, PhD; Dharshan Vummidi, MD

*University of Minnesota, Minneapolis, MN*: Joanne Billings, MD; Abbie Begnaud, MD; Tadashi Allen, MD

*University of Pittsburgh, Pittsburgh, PA*: Frank Sciurba, MD; Jessica Bon, MD; Divay Chandra, MD, MSc; Carl Fuhrman, MD; Joel Weissfeld, MD, MPH

*University of Texas Health, San Antonio, San Antonio, TX*: Antonio Anzueto, MD; Sandra Adams, MD; Diego Maselli-Caceres, MD; Mario E. Ruiz, MD; Harjinder Singh

