## Supplemental Data for "Characterization of a COPD-Associated *NPNT* Functional Splicing Genetic Variant in Human Lung Tissue via Long-Read Sequencing"

**Supplemental Methods**

*Targeted Enrichment and Library Preparation for ONT Long-read RNA Sequencing of LTRC Lung RNA*

RNA was reverse transcribed using Smartscribe RT (Clontech). For each sample, reverse transcription was primed with a different OligodT primer containing 30 Ts, a 10nt sample index, and a universal ISPCR priming site. The reverse transcription reaction also contained a template switch oligo (TSO-Smart-seq2) to attach the same universal priming site to the 5’ end of transcripts. After reverse transcription, RNA and primer dimers were digested using RNAseA and Lambda Exonuclease (NEB) after which cDNA was amplified using the Kapa Biosystems HiFi HotStart ReadyMix (2X) (KAPA) with the following heat-cycling protocol: 37°C for 30 minutes, 95°C for 30 seconds followed by 12 cycles of (98°C 20 seconds; 67°C 15 seconds; 72°C for 6 minutes). The reaction was then purified using SPRI beads at a 0.65:1 ratio (to retain cDNA longer than 500bp) and eluted in H2O. 250ng of the resulting indexed cDNA from each of the ten samples was pooled. The resulting pool of 2.5ug of cDNA was then enriched for NPNT cDNA and a panel of 9 other genes using the xGen Predesigned Gene Capture Pools (Integrated DNA Technologies) according to the manufacturer’s protocol and using blocking oligos. Enriched cDNA was amplified using the Kapa Biosystems HiFi HotStart ReadyMix (2X) (KAPA) with the following heat-cycling protocol: 95°C for 30 seconds followed by 12 cycles of (98°C 20 seconds; 67°C 15 seconds; 72°C for 6 minutes). This cDNA amplification and enrichment was performed in two separate technical replicates.

Enriched re-amplified cDNA was then sequenced on the Oxford Nanopore Technologies (ONT) MinION sequencer using the R2C2 method^1-4^.

In short, 100ng of cDNA is circularized using 100ng of a DNA splint (replicate 1: splint 6, replicate 2: splint 1, 3, and 5) and 2x NEBuilder HiFi DNA Assembly Master Mix (NEB). This mix was incubated at 50C for 60 minutes. Non-circularized cDNA was digested by adding 5ul of NEBuffer 2, 3ul Exonuclease I, 3ul of Exonuclease III, and 3ul of Lambda Exonuclease (all NEB) and adjusting the volume to 50ul using H2O. This reaction was then incubated 37°C for 16hr followed by a heat inactivation step at 80°C for 20 minutes. Circularized DNA was then extracted using SPRI beads with a size cutoff to eliminate DNA <500 bp (0.65 beads:1 sample) and eluted in 40 μL of ultrapure H2O. Circularized DNA was split into four aliquots of 10 μL, and each aliquot was amplified in its own 50-μL reaction containing Phi29 polymerase (NEB) and exonuclease resistant random hexamers (Thermo) [5 μL of 10× Phi29 Buffer, 2.5 μL of 10 uM (each) dNTPs, 2.5 μL random hexamers (10 uM), 10 μL of DNA, 29 μL ultrapure water, 1 μL of Phi29]. Reactions were incubated at 30 °C overnight. T7 Endonuclease was added directly to each reaction which were then incubated at 37°C for 2h with occasional agitation. The debranched DNA was then pooled and concentrated using DNA Clean & Concentrator-5 columns (Zymo Research) and >5kb DNA was then extracted from a 1% DNA agarose gel.

*Short-read RNA sequencing of LTRC Lung RNA*

LTRC short read RNA-seq was performed at the Northwest Genomics Center (NGC) according the following standardized workflow.

Library Production

Total RNA is normalized to 7.5ng/ul in a total volume of 50ul on the Perkin Elmer Janus Workstation (Perkin Elmer, Janus II). Poly-A selection and cDNA synthesis are performed using the TruSeq Stranded mRNA kit as outlined by the manufacturer (Illumina, cat#RS-122-2103). All steps are automated on the Perkin Elmer Sciclone NGSx Workstation to reduce batch to batch variability and to increase sample throughput. Final RNA-seq libraries are quantified using the Quant-it dsDNA High Sensitivity assay, and library insert size distribution is checked using a fragment analyzer (Advanced Analytical; kit ID DNF474). Samples where adapter dimers constitute more than 4% of the electopherogram area are failed prior to sequencing. Technical controls (K562,Thermo Fisher Scientific, cat# AM7832), are compared to expected results to ensure that batch to batch variability is minimized. Successful libraries are normalized to 10nM for submission to sequencing.

Sequencing and Alignment

Barcoded libraries are pooled using liquid handling robotics prior to loading. Massively parallel sequencing-by-synthesis with fluorescently labeled, reversibly terminating nucleotides is carried out on the NovaSeq sequencer. Demultiplexed, unaligned BAM files produced by Picard ExtractIlluminaBarcodes and IlluminaBasecallsToSam are converted to FASTQ format using SamTools bam2fq (v1.4). Sequence read and base quality are checked using the FASTX-toolkit (v0.0.13), and sequences are aligned to GRCh38 with reference transcriptome GENCODE release 29 using STAR (v2.6.1d).

Data Analysis QC

Key expression QC metrics were reviewed for outliers. Marginal outliers in quality are identified using the interquartile ranges (IQR; ~1.5 * IQR) and flagged for further review. PCA plots of meta data are generated using PCA analysis and are colored and visualized using standard PCA plotting packages in Python for detection of confounding effects.

**Supplemental Tables**

Supplemental Table 1. Query of rs34712979 for linked SNPs, evolutionary conservation, and overlap with functional regulatory annotation (Haploreg v 4.1).

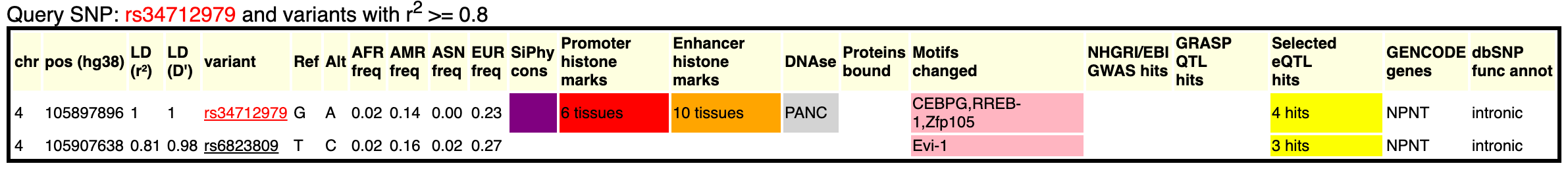

Supplemental Table 2. Significant rs34712979 NPNT leafcutter sQTL results from 49 tissues in GTEx version 8

| Junction Start/End Position | Beta | p-value | GTEx Tissue |
| --- | --- | --- | --- |
| 4:105895723:105897901  (exon 1-2) | -1.05 | 1.61E-48 | Nerve_Tibial |
|  | -0.95 | 3.93E-40 | Adipose_Visceral_Omentum |
|  | -1.12 | 9.83E-40 | Esophagus_Gastroesophageal_Junction |
|  | -0.89 | 1.61E-39 | Adipose_Subcutaneous |
|  | -0.92 | 9.99E-37 | Esophagus_Muscularis |
|  | -1.12 | 3.41E-34 | Pituitary |
|  | -0.91 | 5.64E-30 | Breast_Mammary_Tissue |
|  | -1.10 | 1.26E-26 | Prostate |
|  | -0.65 | 1.99E-20 | Skin_Sun_Exposed_Lower_leg |
|  | -0.58 | 6.52E-15 | Skin_Not_Sun_Exposed_Suprapubic |
|  | -1.13 | 9.27E-11 | Kidney_Cortex |
|  | -0.35 | 1.09E-07 | Muscle_Skeletal |
| 4:105898001:105927336  (exon 2-3) | -0.87 | 5.42E-69 | Thyroid |
|  | -0.62 | 4.02E-38 | Lung |
|  | -0.69 | 2.60E-29 | Artery_Tibial |
| 4:105912238:105927336  (unannotated junction) | 0.73 | 1.04E-24 | Artery_Aorta |
|  | 0.64 | 4.26E-20 | Esophagus_Mucosa |
|  | 0.47 | 1.86E-07 | Colon_Sigmoid |
| Results for rs34712979 where A is the coded/effect allele. | | | |
| Beta is slope from regression model where normalized junctional ratio is the response and dosage of the A allele is the predictor.  p-value is the unadjusted p-value from regression analysis within each tissue | | | |

Supplemental Table 3. Complete rs34712979 lung sQTL results for significant leafcutter clusters.

| Junction | Cluster | p-value | Slope |
| --- | --- | --- | --- |
| 105898001-105902694 | clu_44968 | 0.07 | 0.12 |
| 105898001-105912188 |  | 5.52E-19 | 0.56 |
| 105898001-105927336 |  | 4.02E-38 | -0.62 |
| 105912238-105912543 |  | 0.14 | 0.08 |
| 105912238-105927336 |  | 2.72E-27 | 0.52 |
| 105912651-105927336 |  | 0.01 | 0.14 |
| 105925444-105927336 |  | 0.23 | 0.07 |
| 105927428-105931515 | clu_44969 | 8.51E-11 | 0.31 |
| 105927428-105932614 |  | 0.21 | 0.07 |
| 105927428-105937009 |  | 2.13E-05 | -0.22 |
| 105932703-105937009 |  | 0.00097 | 0.19 |
| 105942702-105958471 | clu_44970 | 0.16 | 0.10 |
| 105942702-105959028 |  | 0.00085 | -0.16 |
| 105958557-105959028 |  | 0.61 | 0.04 |
| 105959126-105967188 | clu_44971 | 0.71 | 0.03 |
| 105959126-105968895 |  | 0.04 | -0.10 |
| 105967444-105968895 |  | 0.72 | 0.03 |
| GTEx lung splice QTL results for rs34712979 and *NPNT* junctions. A is the coded/effect allele. | | | |

Supplementary Table 4: Colocalization posterior probabilities for various combinations of datasets

| **Model** | | | | **Posterior Probability** |
| --- | --- | --- | --- | --- |
| **Datasets sharing unique genetic cause** | **Datasets sharing unique genetic cause** | **Datasets sharing unique genetic cause** | **Datasets sharing unique genetic cause** |  |
| FEV1, FEV1/FVC, COPD, lung-sQTL |  |  |  | 0.9398 |
| FEV1, FEV1/FVC, COPD | lung-sqtl |  |  | 0.0367 |
| FEV1, COPD | FEV1/FVC, lung-sqtl |  |  | 0.0070 |
| FEV1, lung-sQTL | FEV1/FVC, COPD |  |  | 0.0067 |
| FEV1, FEV1/FVC | COPD, lung-sQTL |  |  | 0.0064 |
| FEV1, FEV1/FVC, lung-sQTL | COPD |  |  | 0.0012 |
| FEV1 | FEV1/FVC, COPD, lung-sQTL |  |  | 0.0011 |
| FEV1, COPD, lung-sQTL | FEV1/FVC |  |  | 0.0010 |
| FEV1, COPD | FEV1/FVC | lung-sQTL |  | 2.04E-05 |
| FEV1 | FEV1/FVC, COPD | lung-sQTL |  | 2.02E-05 |
| FEV1, FEV1/FVC | COPD | lung-sQTL |  | 2.00E-05 |
| FEV1 | FEV1/FVC, lung-sQTL | COPD |  | 4.36E-06 |
| FEV1, lung-sQTL | FEV1/FVC | COPD |  | 4.17E-06 |
| FEV1 | FEV1/FVC | COPD, lung-sQTL | lung-sQTL | 4.05E-06 |
| FEV1 | FEV1/FVC | COPD |  | 1.33E-08 |
| FEV1, FEV1/FVC, COPD |  |  |  | 1.78E-39 |
| FEV1, FEV1/FVC, lung-sQTL |  |  |  | 3.63E-42 |
| FEV1, FEV1/FVC | COPD |  |  | 7.91E-43 |
| FEV1 | FEV1/FVC, COPD |  |  | 7.84E-43 |
| FEV1, COPD | FEV1/FVC |  |  | 7.72E-43 |
| FEV1, FEV1/FVC | lung-sQTL |  |  | 3.25E-44 |
| FEV1 | FEV1/FVC, lung-sQTL |  |  | 5.51E-45 |
| FEV1, lung-sQTL | FEV1/FVC |  |  | 5.18E-45 |
| FEV1 | FEV1/FVC | COPD |  | 1.11E-45 |
| FEV1 | FEV1/FVC | lung-sQTL |  | 2.08E-47 |
| FEV1, FEV1/FVC |  |  |  | 1.71E-81 |
| FEV1 | FEV1/FVC |  |  | 5.52E-85 |
| FEV1/FVC, COPD, lung-sQTL |  |  |  | 3.14E-93 |
| FEV1/FVC, COPD | lung-sQTL |  |  | 2.91E-95 |
| FEV1/FVC, lung-sQTL | COPD |  |  | 4.95E-96 |
| FEV1/FVC | COPD, lung-sQTL |  |  | 4.48E-96 |
| FEV1/FVC | COPD | lung-sQTL |  | 1.85E-98 |
| FEV1, COPD, lung-sQTL |  |  |  | 2.66E-126 |
| FEV1, COPD | lung-sQTL |  |  | 2.60E-128 |
| FEV1, lung-sQTL | COPD |  |  | 4.19E-129 |
| FEV1 | COPD, lung-sQTL |  |  | 4.03E-129 |
| FEV1 | COPD | lung-sQTL |  | 1.64E-131 |
| FEV1/FVC, COPD |  |  |  | 1.52E-132 |
| FEV1/FVC, lung-sQTL |  |  |  | 1.39E-134 |
| FEV1/FVC | COPD |  |  | 4.99E-136 |
| FEV1/FVC | lung-sQTL |  |  | 2.34E-137 |
| FEV1, COPD |  |  |  | 1.36E-165 |
| FEV1, lung-sQTL |  |  |  | 1.18E-167 |
| FEV1 | COPD |  |  | 4.54E-169 |
| FEV1 | lung-sQTL |  |  | 2.10E-170 |
| FEV1/FVC |  |  |  | 1.77E-174 |
| FEV1 |  |  |  | 1.55E-207 |
| COPD, lung-sQTL |  |  |  | 1.02E-218 |
| COPD | lung-sQTL |  |  | 1.88E-221 |
| COPD |  |  |  | 1.38E-258 |
| lung-sQTL |  |  |  | 5.01E-260 |
| None |  |  |  | 2.37E-297 |

Supplemental Table 5. SQANTI Isoform Classifications for *NPNT* Long Read Data

| Isoform Alias  (SQUANTI Isoform ID) | Length | Exons** | Associated Transcript | ORF length | Structural Category | Subcategory | Splice Junctions | Contains 3-nt intron retention |
| --- | --- | --- | --- | --- | --- | --- | --- | --- |
| antisense_isoform  (Isoform_15989_5) | 878 | 2 | Novel | 99 | antisense | multi-exon |  |  |
| noncoding_isoform_1  (Isoform_1958_675) | 610 | 4 | Novel | non-coding | novel not in catalog | any annotated donor/acceptor | Canonical |  |
| coding_isoform_1  (Isoform_5261_166) | 4559 | 12 | Novel | 566 | novel not in catalog | any annotated donor/acceptor | non_canonical* | yes |
| noncoding_isoform_2  (Isoform_18081_188) | 1419 | 1 | Novel | non-coding | novel in catalog | mono-exon by intron retention/s | Canonical |  |
| noncoding_isoform_3  (Isoform_21484_261) | 643 | 5 | Novel | non-coding | novel not in catalog | any annotated donor/acceptor | Canonical |  |
| noncoding_isoform_4  (Isoform_4140_256) | 643 | 5 | Novel | non-coding | novel not in catalog | any annotated donor/acceptor | non_canonical* | yes |
| coding_isoform_2  (Isoform_16510_1818) | 1656 | 3 | Novel | 93 | novel in catalog | combination of known junctions | Canonical |  |
| coding_isoform_3  (Isoform_2340_846) | 1663 | 3 | Novel | 94 | novel not in catalog | any annotated donor/acceptor | non_canonical* | yes |
| noncoding_isoform_5  (Isoform_21246_839) | 627 | 4 | Novel | non-coding | novel not in catalog | any annotated donor/acceptor | non_canonical* | yes |
| noncoding_isoform_6  (Isoform_22653_242) | 678 | 5 | Novel | non-coding | novel not in catalog | any annotated donor/acceptor | non_canonical* | yes |
| noncoding_isoform_7  (Isoform_2039_244) | 675 | 5 | Novel | non-coding | novel not in catalog | any annotated donor/acceptor | Canonical |  |
| coding_isoform_4  (Isoform_7467_175) | 1562 | 2 | Novel | 33 | novel in catalog | no combination of known junctions | Canonical |  |
| coding_isoform_5  (Isoform_6461_471) | 4566 | 12 | ENST00000379987.6 | 565 | full-splice match | multi-exon | Canonical |  |
| SQANTI isoform classification of NPNT isoforms identified from long read sequencing.  *Splice site is canonical due to the presence of the minor allele.  **Full length isoforms were defined as those containing all 12 exons, the remaining isoforms were considered short | | | | | | | | |

Supplemental Figure 1: NPNT amino acid sequence analysis

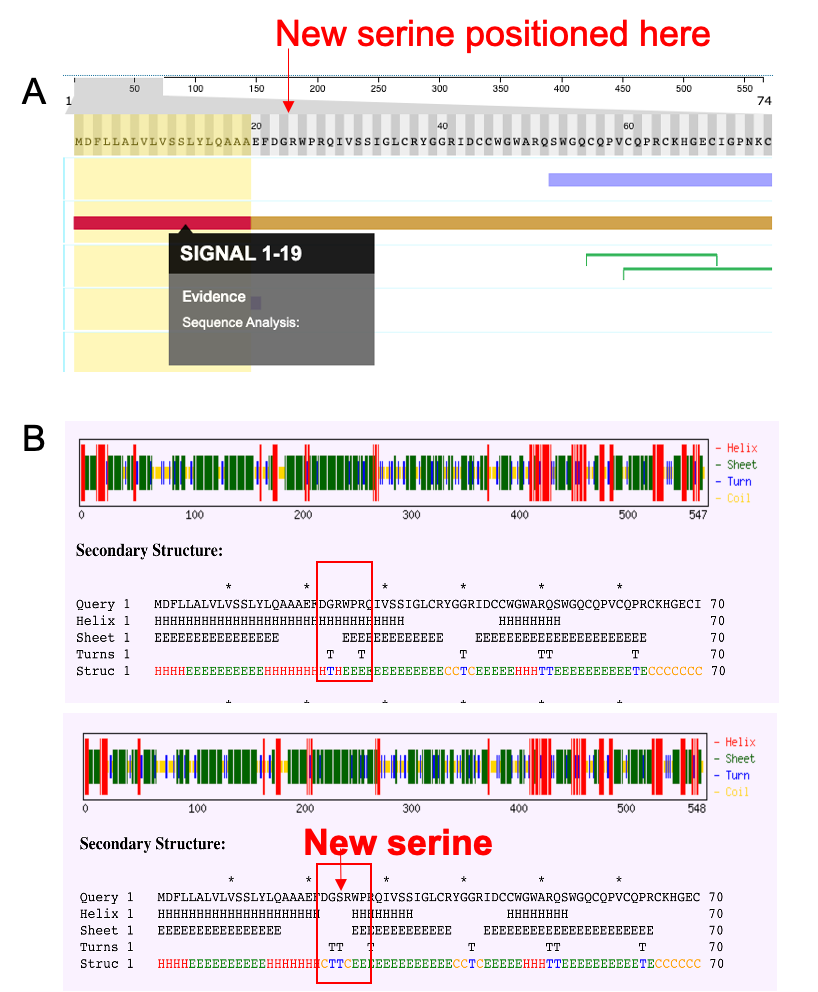
